# Lost in time: temporal monitoring elicits clinical decrements in sustained attention post-stroke

**DOI:** 10.1101/2020.11.30.20239921

**Authors:** MB Brosnan, PM Dockree, S Harty, DJ Pearce, JM Levenstein, CR Gillebert, MA Bellgrove, RG O’Connell, IH Robertson, N Demeyere

## Abstract

**Objective:** Mental fatigue, ‘brain fog’ and difficulties maintaining engagement are commonly reported issues in a range of neurological and psychiatric conditions. Traditional sustained attention tasks commonly measure this capacity as the ability to detect target stimuli based on sensory features in the auditory or visual domains. However, with this approach, discrete target stimuli may exogenously capture attention to aid detection, thereby masking deficits in the ability to endogenously sustain attention over time.

**Method:** To address this, we developed the continuous temporal expectancy test (CTET) where individuals continuously monitor a stream of patterned stimuli alternating at a fixed temporal interval (690ms) and detect an infrequently occurring target stimulus defined by a prolonged temporal duration (1020ms or longer). As such, sensory properties of target and non-target stimuli are perceptually identical and differ only in temporal duration. Using the CTET, we assessed stroke survivors with unilateral right hemisphere damage (*N*=14), a cohort in which sustained attention deficits have been extensively reported.

**Results:** Stroke survivors had overall lower target detection accuracy compared to neurologically-healthy age-matched older controls (N=18). In addition, performance of the stroke survivors was characterised by significantly steeper within-block performance decrements which occurred within short temporal windows (~3 ½ minutes) and were restored by the break periods between blocks.

**Conclusion:** These findings outline a precise measure of the endogenous processes hypothesized to underpin sustained attention deficits following right hemisphere stroke and suggest that continuous temporal monitoring taxes sustained attention process to capture clinical deficits in this capacity over time.

## Introduction

Difficulties concentrating and maintaining engagement, mental fatigue, and ‘brain fog’ are commonly reported in a wide range of psychiatric, neurodevelopmental, and neurological conditions including depression (Deng, Li, Mindfulness, 2014, n.d.), ADHD (Association, 2013) stroke (Rueckert & Grafman, 1996), Alzheimer’s Disease (Perry, Watson, & Hodges, 2000), and, most recently, following COVID-19 (Zhou et al., 2020). Deficits in this fundamental facet of cognition can hinder recovery and exacerbate neurocognitive issues in at least two ways. Firstly, lapses in sustained attention compromise the ability to engage with the task at hand (Smilek, Carriere, & Cheyne, 2010). Therefore, sustained attention deficits limit the extent to which patients can engage with, and accordingly benefit from learning, intervention and rehabilitation programmes (e.g. (Bennett et al., 2002; Blanc-garin, 1994; Robertson, Ridgeway, Greenfield, & Parr, 1997c; van Zandvoort, Kessels, Nys, de Haan, & Kappelle, 2005). Second, the deployment of attention is known to be critical for gating plasticity processes (Polley, Steinberg, & Merzenich, 2006; Recanzone, Schreiner, & Merzenich, 1993). Plasticity processes in turn underlie cortical remapping and functional reorganisation of the brain that are characteristic of recovery (Di Lazzaro et al., 2010; Fu & Zuo, 2011; Hallett, 2001; Phillips, Batten, Tremblay, Aldosary, & Blier, 2015; Qian et al., n.d.). Thus, sustained attention deficits may also hinder the extent to which functional restoration can occur at the neural level.

Several studies in neurologically healthy participants have suggested that sustained attention is supported by a predominantly right-lateralised network (Johannsen et al., 1997; Langner & Eickhoff, 2013; Singh-Curry & Husain, 2009; Warm, Richter, Sprague, Porter, & Schumsky, 1980; Whitehead, 1991), modulated by the locus coeruleus norepinephrine noradrenergic (LC-NE) arousal system (Corbetta & Shulman, 2002; 2011; Posner & Petersen, 1990). In correspondence with findings highlighting a privileged role for the right hemisphere for sustained attention, stroke survivors with damage to RH networks show pronounced deficits in this capacity (Malhotra, Coulthard, & Husain, 2009; Robertson, Manly, Beschin, & Daini, 1997b; Rueckert & Grafman, 1996; 1998; Wilkins, Shallice, & McCarthy, 1987), typically defined by an overall deficit in the ability to detect infrequently occurring visual targets when monitoring a continuous stream of stimuli. Early experimental tasks designed to capture sustained attention typically asked participants to monitor targets over tens of minutes. This would typically elicit performance decrements (so-called time-on-task effects) in healthy individuals (J. F. Mackworth & Taylor, 1963; N. H. Mackworth, 1948; Parasuraman, 1979). Subsequent assessments defined sustained attention as the ability to ‘self-sustain mindful, conscious processing of stimuli’, in order to capture transient lapses in attention while performing more complex, goal-directed tasks of much shorter durations (e.g., the Sustained Attention to Response Test (SART; Manly et al., 2003; Robertson, Manly, Andrade, Baddeley, & Yiend, 1997a). Counter-intuitively, when neurological healthy individuals perform such for protracted periods of time, performance improvements, as opposed to decrements are observed (Brosnan et al., 2018; Staub, Doignon-Camus, Marques-Carneiro, Bacon, & Bonnefond, 2015). Moreover, experimental tasks assessing time on task effects in RH stroke often fail to show performance decrements over relatively short task durations (~8-10minutes; (Malhotra et al., 2009; Rueckert & Grafman, 1996; 1998). This suggests that either RH stroke survivors do not show linear decrements in performance over time on these tasks (which is at odds with anecdotal reports from both RH stroke survivors and their clinicians), or that sustained attention tasks may not be sufficiently sensitive to capture precise deficits in the temporal dynamics of sustained attention difficulties. One limitation of traditional sustained attention tasks is that the discrete onset of sensory target stimuli (i.e., of either visual or auditory targets) may exogenously capture attention and inadvertently ‘support’ sustained attention such that successful performance on the task is not uniquely contingent on endogenous deployment of attentional control. Moreover, the continuous mapping of a transiently-presented target stimulus (e.g., number 3 from a stream of 9 numbers (Manly et al., 2003; Robertson, Manly, Andrade, Baddeley, & Yiend, 1997a) with a specific behaviour (respond or withhold a response), may result in automatic detection which is characteristic of consistent-mapping paradigms (Shiffrin & Schneider, 1977). Thus, these tasks may fail to identify decrements in attentional engagement occurring over relatively shorter time scales.

The current study employed the Continuous Temporal Expectancy Task (CTET; (O’Connell, Dockree, Robertson, Bellgrove, Foxe, & Kelly, 2009a) in which targets, and non-targets differ only in temporal duration and are otherwise perceptually identical. This eliminates any potential for exogenous attentional capture by perceptual features of the stimuli since targets and non-targets differ only in temporal duration. Moreover, previous work has shown that with mind-wandering, time ‘contracts’ (Terhune, Croucher, Marcusson-Clavertz, & Macdonald, 2017), therefore a task necessitating the detection of relatively longer temporal intervals might be particularly sensitive to states of inattention. In the CTET, participants monitor a continuous stream of patterned stimuli that are presented centrally and make a button press when they detect a longer-duration stimulus. This task is sensitive to within block performance decrements (over approximately 3 minute windows) in neurologically healthy individuals and here we assess its potential to elicit performance decrements in RH stroke survivors, a group clinically and experimentally characterised by sustained attention deficits (Malhotra et al., 2009; Molenberghs, Gillebert, Schoofs, & Dupont, 2009; Robertson, Ridgeway, Greenfield, & Parr, 1997c; Rueckert & Grafman, 1996; 1998; Wilkins et al., 1987). We reasoned that the CTET will maximally tax sustained attention in RH stroke survivors and reveal performance decrements over short time windows.

## Methods

### Participants

Fourteen chronic stroke survivors with unilateral damage to the RH were recruited through an existing database of patients (recruited either in acute settings or through self-referrals) interested in research participation at the Cognitive Neuropsychology Centre, Department of Experimental Psychology, Oxford University. All patients were >1 year post stroke (*M*=2.15 years, *SD*=1.07; *note* this data was not available for one self-referred patient). All study participants provided written informed consent, in compliance with relevant protocols approved by the University of Oxford Central University Research Ethics Committee and the experimental procedures were conducted in accordance with the latest version of the Declaration of Helsinki. Eighteen neurologically healthy older adults were recruited through an existing database of healthy older adults interested in participating in research at the Turner Institute for Brain and Mental Health, Monash University, Australia. The experimental protocol was approved by Monash University’s Human Research Ethics Committee before testing and was carried out in accordance with the approved guidelines.

### Demographics

All participants provided their age, gender, and level of education (Table 1). Differences in demographic factors between healthy older adults and the stroke survivors were assessed using one way between subject ANOVAs with ‘Group’ as the between subject factor. There was no evidence to suggest that the stroke survivors and healthy older adults differed in age (F_1,30_=2.99; *p*=.09, Table 1). In an effort to ensure that our results were generalisable to both genders we aspired to obtain an equal number of male and female participants in both groups. Binomial tests conducted separately for the healthy older controls and the stroke survivors indicated that the proportion of male to female participants did not differ significantly from the expected .50 (*p*>.41 for both groups, Table 1). The RH stroke survivors had spent fewer years in formal education (F_1,30_=8.98, *p*=.005, η_p_^2^=.23, Table 1).

**Table 1:** Demographic information for the healthy older adults and RH stroke survivors.

|  | Controls<br>(N=18) | Patients<br>(N=14) |
| --- | --- | --- |
| Age (yrs) | 74.78 (5.99)<br>60-82 | 70.71 (7.32)<br>58-84 |
| Gender | 11 male<br>(47.83%) | 9 male<br>(64.29%) |
| *Education<br>(yrs) | 16.89 (3.45)<br>10-22 | 13.23 (3.40)<br>8-21 |

*Note* Years in education was not available for one stroke participant. \* indicates a significant difference between patients and controls Values denote mean (SD).

### Lesions

Magnetic resonance images (MRI) were acquired on a 3T TIM Trio scanner at the Oxford Centre for Clinical Magnetic Resonance Research (OCMR) for N= 7 patients. These scans were acquired at the chronic stage (i.e., > 6 months post-stroke onset). High-resolution 3D whole-brain T1-weighted MRI scans were acquired using a magnetization-prepared rapid gradient echo sequence (MPRAGE; TR 3000ms, TE 4.7ms, flip angle 8°, 1mm isotropic resolution) and fluid rapid attenuated inversion recovery (FLAIR) images were also obtained acquired (TR 5000ms, TE 397, in-plane resolution 1mm, slice thickness 1.5mm). For the remaining patients (n = 7) clinical imaging was acquired at the acute stage (i.e., < 3 weeks post stroke onset) as part of their hospital admission diagnostics. Of these patients, lesions were delineated using either a clinical T1-weighted MRI (n = 1, voxel resolution 1.78mm x 1.78mm x 1mm, 192 axial slices) or clinical CT (n = 6, slice thickness 5mm, 34.17 (2.40; range 30-36) axial slices). Lesions were manually delineated in native structural space slice-by-slice in the axial plane (MRIcron; McCausland Center for Brain Imaging, Columbia, SC, USA). Lesion masks were smoothed at 5mm full-width at half maximum in the z-direction and binarised using a 0.5 intensity threshold. Native space structural scans and lesion masks were aligned to an MNI equivalent stereotactic space (2mm^3^) specific for each modality and generated from a healthy elderly population. The template spaces and normalization algorithms were performed using “Clinical Toolbox” (Rorden et al., 2012), implemented with Statistical Parametric Mapping 8 (SPM8: Wellcome Department of Cognitive Neuroscience, University College London, UK; http://www.fil.ion.ucl.ac.uk). For two patients, lesions were not yet visible at the time of their clinically acquired acute CT scans. However, both patients’ medical presentation and diagnosis indicated RH stroke. The lesion distributions are shown in Fig. 2A.

**Figure 2.**
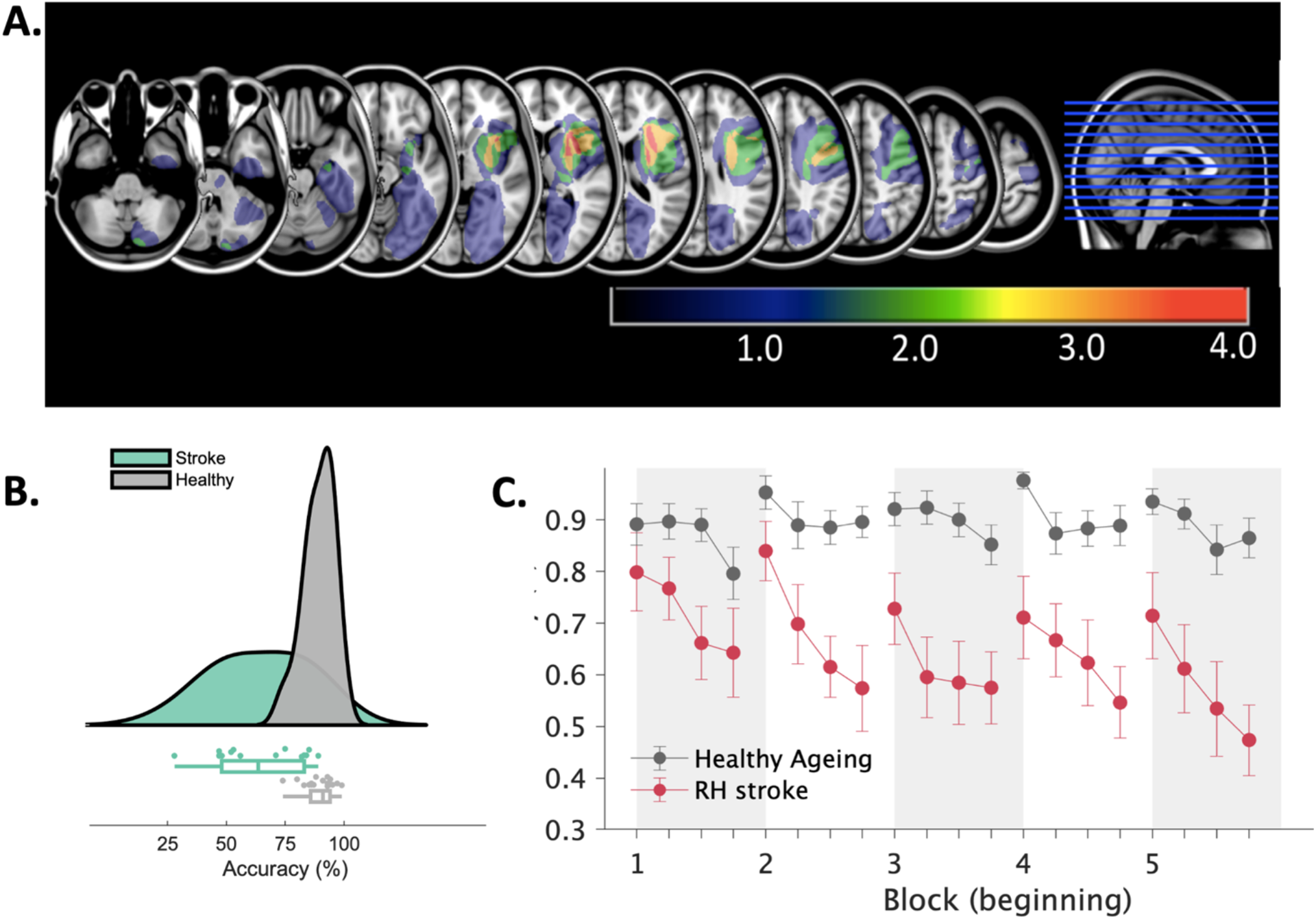
A. Lesion overlay for the right hemisphere stroke survivors. Conjunction maps were created from each patients’ delineated lesion mask (*N*=12) registered to a template space (MNI152 T1 1mm). Intensity values represent the number of subjects with overlapping lesion affected anatomy. **B. Overall accuracy levels for stroke survivors and neurological healthy older adults**. In line with previous sustained attention studies, patients with unilateral RH damage were less accurate on the CTET as compared with neurologically healthy older adults. **C. Performance decrements over time on the CTET**. Performance decrements within the duration of the task blocks were significantly steeper for the patient cohort relative to the neurologically healthy older adults. *Note* alternating grey and white columns depict the start of each task block

### Assessment

#### The Continuous Temporal Expectancy Task (CTET)

The Continuous Temporal Expectancy Task (O’Connell, Dockree, Robertson, Bellgrove, Foxe, & Kelly, 2009b) was employed to measure sustained attention. This is a temporal judgment task designed to elicit frequent lapses of attention. In this task, a patterned stimulus was presented centrally on a grey background and was constantly rotated at 90° angles (see Fig. 1). The pattern stimulus consisted of a single 8 cm^2^ large square divided into a 10 x 10 grid of identical square tiles (0.8 mm^2^), each one diagonally split into black and white halves. The tile orientation shifted by 90° in a random direction (clockwise or counter-clockwise) on each frame change yielding four distinct patterns. In the ‘standard’ trials (~90% of trials) the stimuli were presented for a temporal duration of 690 ms. The participant’s task was to identify the infrequent ‘target’ trials by a button press using their right index finger, where the stimulus was presented for a longer temporal duration. Stimuli were pseudo-randomly presented such that there were between 7 and 15 (average of 11) standard trials between each target presentation.

**Figure 1.**
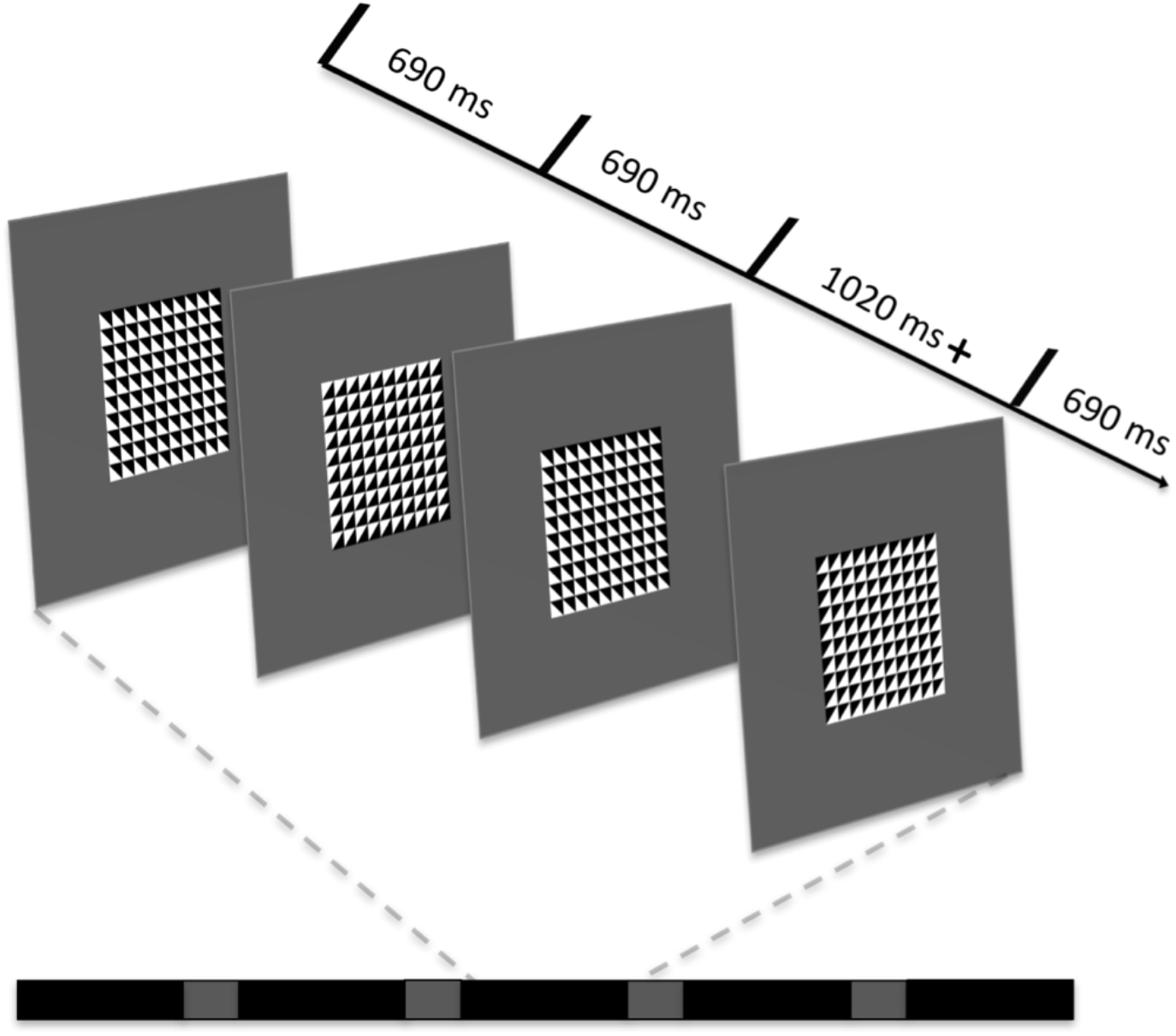
The Continuous Temporal Expectancy Task (CTET). Participants monitored an alternating patterned stimulus. The visual stimulus usually alternated at a fixed temporal interval (690ms) and the participants’ task was to identify stimuli that were presented for a longer interval (1020ms), thereby necessitating target detection in the temporal as opposed to visual domain.

The CTET was designed such that temporal judgments were perceptually undemanding for participants but challenging when tasked to continuously perform the judgments over sustained periods of time. Prior to the experimental blocks, a practice block was administered, where all participants were required to demonstrate 100% accuracy, before advancing to the experimental blocks. For the practice block, three targets were randomly interspersed among 25 standard stimuli. Target stimuli were presented at the target duration of 1020 ms, i.e., 330 ms /47.83% longer than standard trials. If participants missed one or more target stimuli, the block was performed again. If the participant still failed to identify all the targets, the duration of the target stimulus was adapted such that all participants demonstrated 100% accuracy on two consecutive blocks. For the healthy older adults, all participants identified the target stimuli at 100% accuracy during the practice blocks at the target duration of 1020 ms. Of the stroke survivors, 7 identified the target stimulus at a duration of 1020 ms at 100% accuracy. For the other 7 patients, the duration of the target stimulus was adapted as follows such that all participants demonstrated 100% accuracy on two consecutive practice blocks: If the patient did not achieve 100% accuracy during two consecutive blocks within the first three practice blocks, the duration of the target stimulus was increased to 1140ms. Participants were then given two practice blocks with this target duration. If 100% accuracy was achieved during these blocks, then this target duration was set at 1140ms for the rest of the experiment. If the patient did not exhibit 100% accuracy at this duration, the target duration was subsequently increased again to 1500ms, and following this to 1800ms. In order to ensure the stroke survivors fully understood the task requirements all patients were required to complete two consecutive practice blocks at 100% accuracy before proceeding to the experimental trials. For the experimental trials, the target durations for the stroke survivors were either 1020ms (*N*=7), 1140ms (*N*=2), 1500ms (*N*=4), or 1800ms (*N*=1).

This tailored approach was adopted to ensure that any observed differences in attention could not be attributed to the perceptual demands of the task. Participants completed 5 blocks of the task and were given a break (~ 1 minute) in between all blocks. Each block consisted of 225 stimulus rotations with a total duration of between 3 min and 5 s and 3 min and 30 s. The number of targets varied between 18 and 22 per block.

### Analyses

Sustained attention was expressed as the percentage of correct target detections. A target was considered correctly identified if a participant responded within 2.07 seconds (i.e. three standard trials) of a target trial. One stroke patient presented with working memory problems, identified by a previous study in the centre using the Oxford Cognitive Screen (Demeyere, Riddoch, Slavkova, Bickerton, & Humphreys, 2015). In an effort to exclude the possibility that these deficits were contributing to performance on the task, this patient was required to recall the task instructions at the end of each block. Outliers were defined as participants whose overall accuracy levels were greater than 3 times the interquartile range (IQR), which was determined separately for the healthy older adults and stroke survivors.

We examined overall accuracy across all blocks of the task (Fig. 2B), and we also evaluated changes in accuracy both within each block and across blocks (Fig. 2C). Differences in accuracy between the stroke survivors and healthy older adults were evaluated using a repeated-measures mixed ANOVA with ‘Performance Decrement Within Blocks’ (within four quartiles of each block) and ‘Performance Decrement Across Blocks’ (five blocks) as within subject factors. ‘Group’ (RH stroke, healthy ageing) was included as a between subject factor. In order to assess the putatively restorative effect of task break on performance, the average of the 4^th^ quartile computed across blocks 1, 2, 3, and 4 was compared against the average of the 1^st^ quartile across blocks 2, 3, 4, and 5 using a repeated-measures ANOVA with break as a within-subjects, and group as a between-subjects factor.

In order to rule out the possibility that differences in formal education between the two groups could be associated with any observed effects, all significant ANOVAs were repeated years in formal education included as a covariate.

## Results

### Overall Target Detection (Accuracy)

The rate of false alarms (i.e., the number of button presses in the absense of a target stimulus) was low on the CTET (*M*=3.36, *SD*=2.03, range = 1.2-9.4%) indicating that all participants were performing well above chance level and had correctly understood the instructions. In line with previous reports investigating overall performance levels in RH stroke survivors (Robertson, Manly, Andrade, Baddeley, & Yiend, 1997a; Rueckert & Grafman, 1996; 1998), a main effect of Group (F_1,29_=25.34, *p*<.0005, η_p_^2^=.47, Fig. 2B; RH stroke M(SD) 64.27% (19.17); Healthy older controls M(SD) 89.29% (6.56)) indicated that the patients were significantly less accurate at identifying target stimuli, compared with the healthy older controls. This main effect remained significant after controlling for years of education (F_1,28_=16.70, *p*<.0005, η_p_^2^=.37).

### Time on Task Decrements

#### Performance Decrement Within Blocks

A main effect of Quartile (F_3,87_=15.58, *p*<.0005, η_p_^2^=.35) demonstrated that accuracy on the task decreased within each block (see Fig. 2C), as confirmed by a significant linear contrast (*F*_1,29_=42.36, *p*<.0005, *η*_p_^2^=.59). Critically, this was qualified by a Group **X** Quartile interaction (*F*_3,87_=3.21, *p*=.03, *η*_p_^2^=.10). Follow up repeated-measures ANOVAs with Quartile as the within-subjects factor were conducted separately for the neurologically healthy older group and the RH stroke survivors. For both the healthy group (*F*_3,48_=3.58, *p*=.02, *η*_p_^2^=.18) and the stroke survivors (*F*_3,39_=11.19, *p*<.0005, *η*_p_^2^=.46), there was a main effect of Quartile, best captured by a linear fit (healthy; *F*_1,16_=7.41, *p*=.02, *η*_p_^2^=.32; stroke: *F*_1,13_=38.37, *p*<.0005, *η*_p_^2^=.75). Thus, the significant Group **X** Quartile interaction was driven by a significantly steeper within-block decrement in the accuracy of the RH stroke survivors relative to the healthy older controls. When controlling for years of education the main effect of Quartile was no longer significant (F_3,84_=1.14, *p*=.34). Crucially however the Group x Quartile interaction remained significant (F_3,84_=3.73, *p*=.01, *η*_p_^2^=.12).

### Time on Task Decrements

#### Performance Decrement Across Blocks

There was no main effect of Block (F_4,116_=1.78, p=.14) but there was a Group x Block interaction (F_4,116_=2.75, p=.03, η_p_^2^=.09) which remained significant after controlling for education (F_4,112_=2.66, p=.04, η_p_^2^=.09) indicating that the two groups differed in the way their performance changed across blocks. Follow up repeated measures analyses conducted for each group separately failed to resolve the source of this interaction with no main effect of Block for either the stroke (F_4,52_=2.31, p=.07, η_p_^2^=.15) nor the healthy older cohort (F_4,64_=.85, p=.5). As such, these results suggest that while the stroke survivors exhibit particularly sharp decrements in their ability to sustain attention within blocks, we found no evidence that the stroke survivors differed from controls in their capacity to sustain attentional engagement over coarser timescales (i.e., across blocks). Finally, there were no other significant interaction terms (Quartile x Block, and Quartile x Block x Group both *p*>.72).

### Break periods to restore performance

The findings thus far indicate that deficits in attention arise from deficits in the capacity to maintain engagement over short time periods (within block), while performance over long time periods (across blocks) remained stable. This suggests that the short break periods in between blocks exerted a restorative effect over performance. Formal analyses supported this hypothesis. A main effect of Break was observed (F_1,29_=36.76, p<.0005, η_p_^2^=.56), and there was no significant Break x Group interaction effect (F_1,29_=3.30, p=.08, η_p_^2^=.10). As such, for both the healthy older adults and the RH stroke survivors, performance during the fourth quartiles (healthy older *M*=85.83%, *SD*=12.07, RH stroke *M*=58.45%, *SD*=23.78) was significantly lower than during the first quartile (healthy older *M*=94.63%, *SD*=7.68; RH stroke *M*=74.79%, *SD*=21.23), thus demonstrating that irrespective of group, the performance deficit was offset by rest periods between blocks.

## Discussion

Here we show that monitoring attention in the temporal domain captures clinical deficits in maintaining attention within a shorter time window (a linear decline in performance within ~ 3 ½ minutes) than previously noted with RH stroke survivors (Robertson, Ridgeway, Greenfield, & Parr, 1997c; Rueckert & Grafman, 1996; 1998). Our results indicate that well-established clinical deficits in sustained attention (overall target detection accuracy) following RH stroke (Robertson, Ridgeway, Greenfield, & Parr, 1997c; Rueckert & Grafman, 1996; 1998) may arise from a specific impairment in the ability to maintain engagement over relatively short temporal windows. As such, these results suggest that the CTET (O’Connell, Dockree, Robertson, Bellgrove, Foxe, & Kelly, 2009a) is a sensitive tool for capturing and examining performance decrements in sustained attention over short temporal windows.

There are several aspects of the CTET that make it appealing for assessing attentional deficits, particularly in patient populations. Firstly, the target stimulus is adapted such that 100% accuracy is exhibited during the practice trials preceding the experimental blocks. Thus, the task is not confounded by an overall perceptual deficit in the stroke survivors. Secondly, unlike many sustained attention tasks (e.g. (Malhotra et al., 2009; Manly et al., 2003; Rueckert & Grafman, 1996; 1998)), identification of the target in the CTET occurs in the temporal domain rather than being based on perceptual characteristics of targets and non-targets. This is potentially very advantageous in producing a performance decrement as visual search processes which may help with the identification of target stimuli, such as stimulus-driven attentional capture and stimulus-response mapping, are reduced in the CTET, thus placing greater demands on the continuous monitoring process and taxing the sustained attention system.

Despite the stroke survivors demonstrating steep within-block performance declines, they also exhibited the capacity to temporarily revive their sustained attention following the brief within-block rest periods, an observation supported by significantly higher performance during the first quartiles of blocks 2,3,4, and 5, as compared with the last quartiles of blocks 1,2,3 and 4. In the current study, the within-block performance decrement observed in the RH stroke survivors did not get worse with time (i.e., there was no interaction between this effect and the overall performance decrement over the course of the experiment). This suggests that the sustained attention system can be revived within intervals as short as the between block breaks of approximately one minute. This is in line with literature suggesting the sustained attention system can be manipulated by increasing levels of alertness through the voluntary control over biophysiological markers of arousal (O’Connell, Bellgrove, Dockree, & Lau, 2008; Robertson, Tegnér, Tham, & Lo, 1995), or by presenting salient sensory stimuli which act as external alerting cues (Finke, Matthias, Keller, Müller, & Schneider, 2012; Robertson, Mattingley, Rorden, & Driver, 1998).

In this study we observed large interindividual differences in sustained attention capacity on the CTET, particularly in the patient cohort. An avenue for future work will be to highlight neural sources of variability on the CTET along with the functional, neuropsychological, and clinical implications of deficits in the capacity to sustain engagement. While our study highlights the feasibility of the CTET to effectively elicit sustained attention deficits in a relatively small cohort that is characterised by known difficulties in this capacity, future confirmation of our results will be required in larger-scale clinical studies. In particular, it will be important to assess whether the CTET is sensitive to attention deficits following exclusively right hemisphere infarcts or whether this task is sensitive to other neurological events, including brain stem damage, left hemisphere lesions, traumatic brain injury, and neuroinflammation. Moreover, our groups differed both in terms of geographical location and levels of educational attainment and future research should address whether cultural, environmental, and cognitively enriching factors including education, may mitigate deficits in the capacity to sustain engagement.

One likely explanation for these sharp decreases in arousal following RH damage is dysfunction of the noradrenergic system. Noradrenaline (norepinephrine), a neurotransmitter emanating from the locus coeruleus with widespread projections throughout the cortex, plays a well-established role in maintaining arousal (Corbetta & Shulman, 2002; 2011; Posner & Petersen, 1990) for a review see (Sara, 2009). RH dominance of noradrenaline is evident from asymmetric organisation of the noradrenergic system in rats (Robinson, 1979; Robinson & Coyle, 1980), higher noradrenergic concentrations in the right thalamus in post-mortem human neurochemical studies (Debecker, Laget, & Raimbault, 1978), and increased BOLD activation exclusively within the RH during pharmacological upregulation of noradrenaline (Grefkes, Wang, Eickhoff, & Fink, 2010). Pharmacological interventions such as noradrenergic agonists and methylphenidate that modulate noradrenergic levels within prefrontal regions have been shown to improve sustained attention performance on the CTET in healthy younger adults (Dockree et al., 2017), and preliminary evidence suggests that this may be a useful approach to improve sustained attention deficits associated with damage to the RH (Malhotra, Parton, Greenwood, & Husain, 2006; Singh-Curry, Malhotra, Farmer, & Husain, 2011). Future work should directly explore whether noradrenergic dysfunction underlies the pronounced decrements in sustained attention observed in the current study, and whether manipulating activity within this right-lateralised system, for example through pharmacological interventions (Dockree et al., 2017), behavioural training paradigms (Milewski-Lopez et al., 2014), fMRI neurofeedback training techniques (deBettencourt, Cohen, Lee, Norman, & Turk-Browne, 2015; Habes et al., 2016), or brain stimulation (Brosnan et al., 2018; 2017) might ameliorate these performance decrements.

Finally, although specific functional impairments resulting from a stroke have often been considered in the context of modular damage (e.g. (Broca, 1865), there is a growing consensus that it is not only damage to specific regions but rather disruptions to inter-connected neural networks which are associated with functional impairments (e.g. (Forkel et al., 2014) cf (Baldassarre, Ramsey, Siegel, Shulman, & Corbetta, 2016; Corbetta & Shulman, 2011). With regards to the sustained attention system, recent multivariate analyses of fMRI data have identified a widespread network of cortical, sub-cortical and cerebellar regions underlying the capacity to maintain attentional engagement (deBettencourt et al., 2015; Rosenberg et al., 2016). Rosenberg and colleagues recently showed that only 27% of a multivariate pattern analysis (MVPA)-derived network which successfully predicted the ‘high’ attention state both in healthy younger adults, and children and adults with attentional deficits (ADHD symptoms) included prefrontal and parietal nodes. These findings do not discount fronto-parietal networks contributions to maintaining sustained attention (Rueckert & Grafman, 1996; 1998; Wilkins et al., 1987) but rather demonstrate the role of these regions within a widespread network (Shalev, Humphreys, & Demeyere, 2016). The heterogeneity of the lesions in the current cohort support these findings and suggest damage to widespread cortical regions within the right-lateralised noradrenergic-network results in deficits maintaining sustained attention over time (cf, (Corbetta & Shulman, 2011).

The current findings hold important translational implication for informing cognitive assessment and rehabilitation strategies for individuals who have experienced RH stroke, and in particular, they speak to the need for employing assays that are sensitive to detecting deficits over short time intervals when appraising the integrity of sustained attention in this cohort.

## Data Availability

Data supporting the findings of this study are available from the corresponding author upon request.

## Acknowledgements

We would like to sincerely thank the late Prof Glyn Humphreys for his valuable contributions to the design and implementation of this project. We wish to thank Tara Anne Maguire for her assistance with data collection, and our participants in Melbourne and Oxford for generously volunteering their time. This work was supported by the European Union FP7 Marie Curie Initial Training Network Individualised Diagnostics & Rehabilitation of Attention Disorders (grant number 606901), a Marie Skłodowska-Curie Fellowship from the European Commission (AGEING PLASTICITY; grant number 844246), and supported by the NIHR Oxford Health Biomedical Research Centre. The Wellcome Centre for Integrative Neuroimaging is supported by core funding from the Wellcome Trust (203139/Z/16/Z). Paul Dockree was supported by an Irish Research Council Laureate grant: IRCLA/2017/306. The authors declare no conflicts of interest affecting this manuscript.

